# Role of cytokines and other prophetic variables in the development and progression of disease in patients suffering from COVID-19

**DOI:** 10.1101/2020.10.28.20221408

**Authors:** Arif Malik, Saima Iqbal, Sulayman Waquar, Muhammad Mansoor Hafeez

**Affiliations:** Institute of Molecular Biology and Biotechnology (IMBB), The University of Lahore, Lahore-Pakistan

**Keywords:** COVID-19, Cytokine Storm, spike (S) protein, Interleukins, TNF-α

## Abstract

**INTRODUCTION:** Outbreak of the novel COVID-19 infection identifies both causative agents that threaten global pandemic in 2003 and 2011. It is an enveloped virus with spike (S) protein attached that facilitates its receptor binding on the surface. Although it has brought about the global interest for the researchers and medical practitioner in the identification of potential targets that may be addressed in order to cope up with the situation. In the current study potential role of cytokines and related inflammatory markers have been identified that interplays in the progression of disease in COVID-19 patients.

**MATERIALS AND METHODS:** Current study substitutes hundred and fifty (n=150) patients with novel-COVID-19 and hundred (n=100) healthy controls. After getting informed consent serum samples of the participants were collected and analyzed for their significance in the disease progression. Levels of Interleukins i.e., (IL- 1,6,8,10,11) and tumor necrosis factor-alpha (TNF-α) were determined with help of their specific spectrophotometric and ELISA methods.

**RESULTS:** Findings of study show significant increase in the levels of interleukins and TNF-α that signifies the presence of “cytokine storm” in worsening the condition in respect to the exposure of COVID-19. Levels of IL-1 and 6 were significantly higher in patients (98.69±39.35pg/ml and 71.95±28.41 pg/ml) as compared to controls (30.06±14.19pg/ml and 9.46±3.43pg/ml) where, (p=0.001 and 0.007). It also suggests that IL-6 is most sensitive test with about (98%) sensitivity in comparison with 96%,95%, 95%,93% and 92% in case of IL-10,1,8,11 and TNF-α respectively.

**CONCLUSION:** Current study elucidate the effects of cytokines and respective inflammatory markers in the progression of the COVID-19. Findings show that activation of macrophages and neutrophils have significant role in the worsening of the symptoms and progression of the viral infection. Thus, use of certain blockers in initial stages could serve with potent benefits in coping up the infectious condition.

## INTRODUCTION

Recent emergence of the novel virus and stated pandemic named COVID-19 is raging all over the world and is significantly responsible for the threat to health-care systems and to the lifestyles of the individuals living worldwide [1]. Once after the spread of the pandemic identification of the virus and its strains is very important. Traditional approaches that may be used for the identification of the viruses include culturing of the virus, staining of the antigen, electron microscopy and its serology [2]. Apart from all the techniques that can be used in determining the characteristics of the onset disease there are some other standardized procedures that need to be characterized over time to identify and state agents involved in both chronic and acute infections. Corona viruses (CoVs) belong to the diverse zoonotic group of viruses that were identified in different animals that is responsible for their unique characteristics and thus serve as the reservoir of corona virus prevalence and thus their expression increased as the population increased in the undeveloped regions of the world. These are niched as enveloped viruses having Ribonucleic Acid (RNA) nature. Onset of the virus is usually responsible for the infections that involves different enteric or respiratory disorders. Whereas, before the outbreak of SARS-CoV the virus was believed only to cause mild fever that may be referred as “colds” and rarely was observed to cause infection of lower respiratory tract [3]. Although the studies suggest that there is high prevalence of the disease in the elder people especially with increased cardiovascular risks including hypertension. There are some interesting facts that hints on pulmonary edema, dry cough as the distinguishing features of the disease progression along with the increased inflammation at cellular level [4]. Thus, here arises the need of significant procedures to overcome the said characteristics of the disease to manage the status of the patients accordingly. The spike (S) protein attached with RNA single stranded virus helps the entry of the virus in to the cell. Entry is specifically dependent upon the affinity of the said protein with the surface receptor. In addition to the said mechanism the entry of the virus or binding of the protein and receptor requires priming of the spike protein that cleaves it into two subunits namely S1 and S2 which facilitates the fusion of cellular and viral membranes [5]. Along the angiotensin converting enzyme-2 receptor another cellular protease is mandatory that is known as TMPRSS2 for the fusion of viral membrane. Although initially the pathogenesis of the coronavirus was not known although with the help of ongoing investigations it was believed that COVID-19 was specific respiratory disease that only effects lungs. Literature and trials carried out executed that primary node of the said infection was close human-human contact that enables the tiny droplets to reach from infected to uninfected individual [6]. Thus the rapid spread and potential of one infected person to spread the disease was the major reason behind the fatality of the disease [7]. Findings of the studies suggest that cytokine storm may initiate viral sepsis and inflammatory-induced lungs injury that contributes in the progression of other complications like respiratory failure, septic shock, organ failure and potentially death [8]. Outcomes from the data collected from infected individuals show spontaneous increase in the levels of IL-6 and c- reactive proteins (CRP) that signifies the potential for the use of anti-inflammatory agents to cope up with the effects of corona virus. The present study strives to bring the potential role of cytokines and related factors that interplay in the progression of infection in patients with COVID-19.

## MATERIALS AND METHODS

For the current study one hundred and fifty (n=150) patients with clinically confirmed COVID-19 and hundred (n=100) healthy controls were substituted. All of the participants were then provided with informed consents and required information were acquired accordingly. Five (5ml) of the blood was drawn from each of the participant and their serum was separated and stored at −80°C for their future analysis. The study was approved from the Departmental Research Committee (DRC), Institute of Molecular Biology and Biotechnology (IMBB), The University of Lahore. Samples were subjected to evaluation of their inflammatory markers i.e., Interleukins (IL-1B,6,8,10,11 and Tumor Necrosis Factor-alpha (TNF-α) with the help of their respective ELISA kits (abcam USA). Results were then subjected to their statistical analysis using SPSS v.21. Independent T-test was carried out and the results were expressed in Mean ± S.D where p<0.05 remained as significant. Moreover, Pearson correlation, NPV, PPV, Sensitivity and Specificity of said variables were carried out.

## RESULTS

Findings of the study suggest and correlates the significance of the evaluated markers in the patients with COVID-19 and in healthy controls. Levels of determined interleukins i.e., IL- 1,6,8,10 and 11 were significantly increased in patients (98.69±39.35pg/ml, 71.95±28.41pg/ml, 81.54±29.59 pg/ml, 14.19±5.43pg/ml and 32.25±11.5pg/ml) as compared to controls (30.06±14.19pg/ml, 9.46±3.43pg/ml, 46.56±17.63pg/ml, 5.21±2.04pg/ml and 22.86±7.61 p= 0.001, 0.007, 0.003, 0.008 and 0.013) respectively. Likewise, marker for the septic shock TNF-α was also significantly increased in COVID-19 patients (0.554±0.21 pg/ml) as compared to control (8.12±2.91) as shown in Table 01.

**Table 1:**
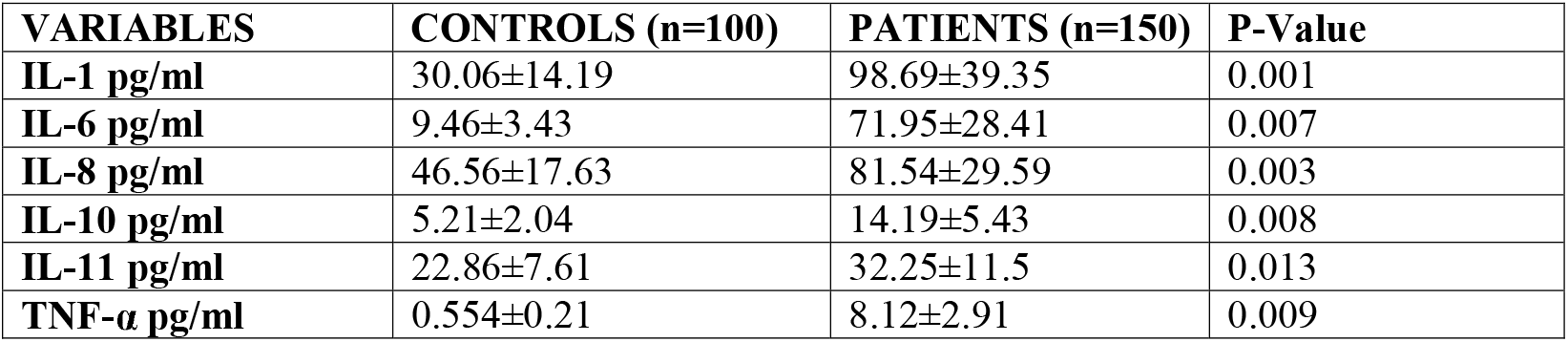
Levels of Different Prophetic Variables in the COVID-19 patients.

Moreover, the findings of interleukins and TNF-α were strived to find out the specificity and sensitivity of the variables along with they were subjected to prediction of positive predictive variables (PPV) and negative predictive variables (NPV). Results demonstrate IL-6 to be most sensitive and specific markers. Sensitivity of the interleukins remained (95%, 98%, 95%, 96%, 93% and 92%) for IL1,6,8,10,11 and TNF-α respectively as shown in Table 2. Table also shows the levels of specificity (SP), PPV and NPV remained (85%,89% and 95% for IL1, 87%,90% and 97% for IL-6, 70%,89%,80% for IL-8, 76%,88%,85% for IL10, 76%,84%,88% for IL-11 and 78%,86%,87% for TNF-α.

**Table 2:**
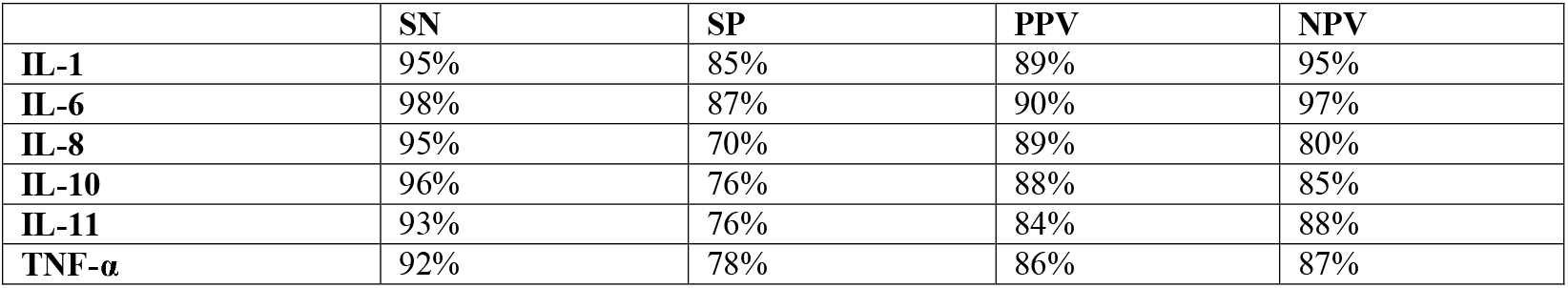
Contingency table showing Sensitivity (SN), Specificity (SP), Positive predictive variable (PPV) AND Negative predictive variable (NPV) OF IL1, 6,8,10,11 and TNF-α.

## DISCUSSION

Progression of the acute respiratory diseases under the influence of the COVID-19 RNA virus, it shows that the patients exposed to the virus have increased inflammatory markers along as the disease progresses in them. It proposes as already stated in the literature it has specific binding affinity with its receptor that mainly includes angiotensin converting enzyme-2 receptor (ACE-2R) along with the cellular protease known as TMPRSS2 [9]. The said receptors are most evidently found in the alveoli of lungs. Apoptotic pneumocyte type II cells release certain molecular patterns [damage associated molecular patterns (DAMPs) and pathogen associated molecular patterns (PAMPs)], inter and intracellularly [10]. Once the RNA virus binds the receptor, complex is taken up by the endocytic cells that is responsible for the initiation of different cascades including the activation of immune response of the cells including recruitment of neutrophils, activation of macrophages and formation of inflammatory complexes that in-turn are responsible for the activation of the certain caspases. The active caspases then convert pro- interleukins to their active forms [11] and thus, leads to the cytokine storming of the cell. Thus, it brings the massive production of inflammatory cytokines commonly known as (cytokine storm) that remains associated with the immune-pathogenesis and elevated rate of mortality upon the onset of the corona virus. Another factor NFkB (transcription factor nuclear factor kappa-light chain enhancer of activated B cells) remains a major regulator of the inflammatory and immune response that comes to rise as the response of the cells [12]. It may be well defined that due to the early induction of NFkB in the scenario induces the over production and over expression of Interleukins i.e., IL6, IL8 and Tumor Necrosis Factor-alpha (TNF-α) [13]. It can be assumed by the findings of current study that levels of said interleukins (IL-1,6,8,10,11 and TNF-α) were significantly exaggerated in patients of COVID-19, if compared with controls as shown in Figure 01 and Table 3 (IL-1 Vs IL-6, r=0.795**), (IL-1 Vs TNF-α, r=0.701**) and (IL-6 Vs TNF-α, r=0.633**). The concurrent cytokine storm may also be involved in the production of pro- coagulants and decreased anti-coagulants that is an important insight for the disseminate vascular coagulation (D.I.C) [14]. As the virus is self-replicated after binding with ACE-2 receptor in pneumocyte type II cells [15] in the alveolus it gets dispersed intra and extra-cellular that leads to the apoptotic type I and II pneumocyte cells. Collectively they are responsible for enhanced levels of antiviral peptides, decreased levels of surfactants, alveolar collapse, hypoxemia, hypercabia, release of leukotrienes and prostaglandins that uptakes the inflammatory stimuli and generates the symptoms of the viral infection including dry cough, and hyperthermia [16]. Findings of the study show that onset of the viral infection not only leads to the altered immune response but it also affects the central nervous condition by the release of certain leukotrienes that are responsible for the induced symptoms of the disease.

**FIG 1:**
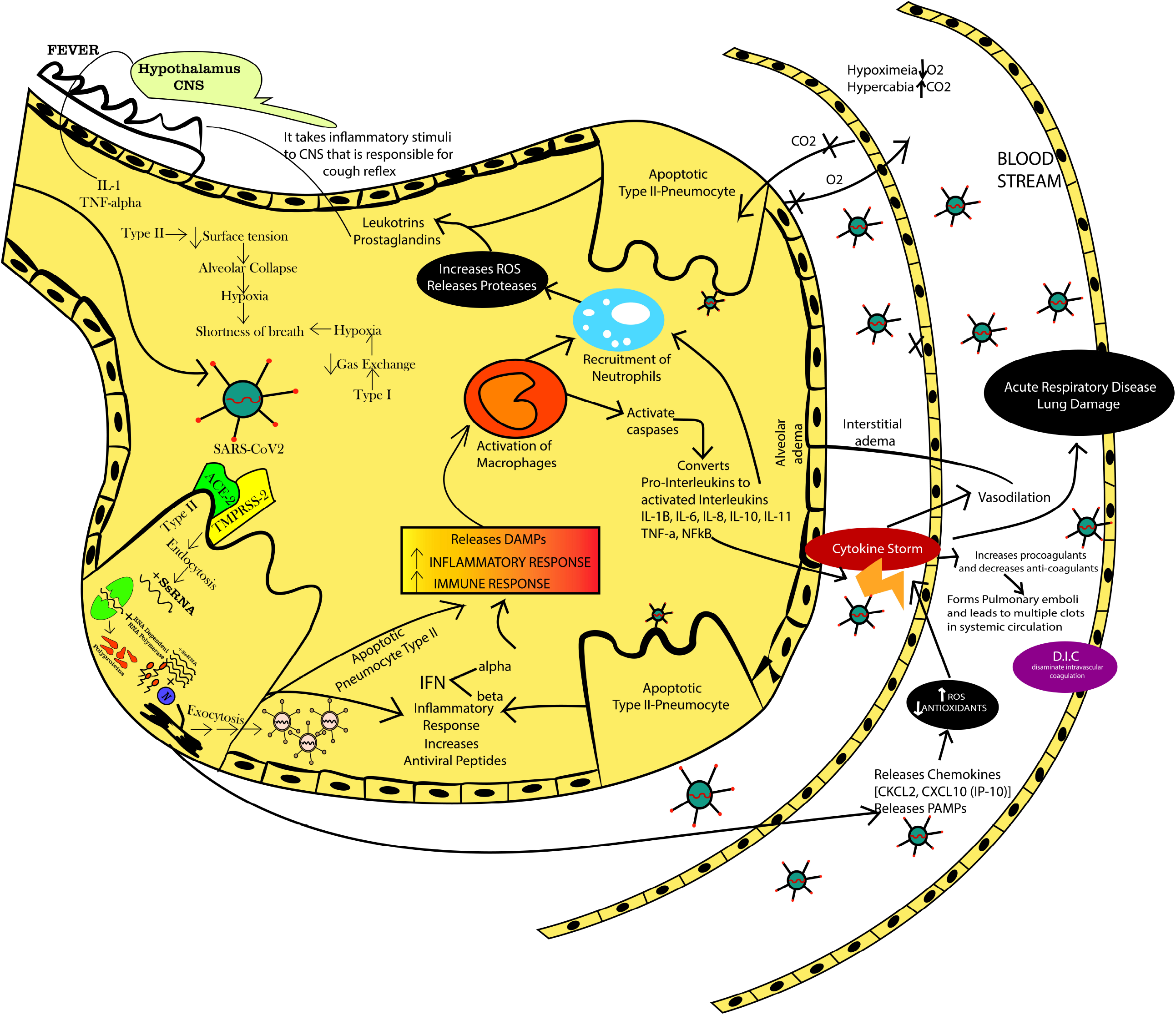
Molecular Mechanism of Interleukins and pro-inflammatory cytokines involved in the progression of COVID-19.

**Table 3:**
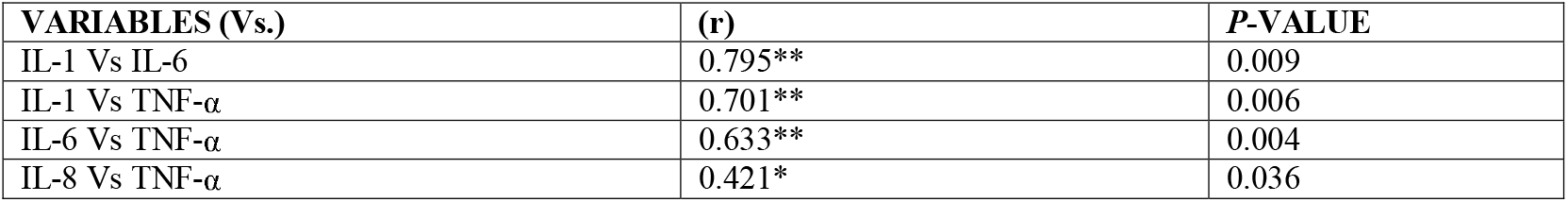
Pearson Correlation Table among the variables estimated in the COVID-19 patients.

## CONCLUSION

Present study demonstrates the potent effects of the cytokine and pro-inflammatory markers in the onset of the viral infection. As after the successful self-replication of the virus, it initiates certain cascades that leads to the apoptosis of the pneumocyte cells and upregulates certain peptides and proteases that are responsible for the regulation of immune response in the alveolar cells and ultimately to the cytokine storm. Thus certain targets that could intercept with the conventional activation of cytokines, macrophages, neutrophils could potentially help in ruling out certain infectious outcomes. Therefore, use of specific blockers for the interleukins or neutralizing the effects of these cytokines can help in coping up with the infectious condition.

## Data Availability

Required related DATA will be furnished on demand

